# Emergency department care transition barriers: a qualitative study of care partners of older adults with cognitive impairment

**DOI:** 10.1101/2022.06.12.22276302

**Authors:** Cameron J. Gettel, Peter T. Serina, Ivie Uzamere, Kizzy Hernandez-Bigos, Arjun K. Venkatesh, Andrew B. Cohen, Joan K. Monin, Shelli L. Feder, Terri R. Fried, Ula Hwang

## Abstract

**INTRODUCTION:** After emergency department (ED) discharge, persons living with cognitive impairment (PLWCI) and their care partners are particularly at risk for adverse outcomes. We sought to identify the barriers experienced by care partners of PLWCI during ED discharge care transitions.

**METHODS:** We conducted a qualitative study of 25 care partners of PLWCI discharged from four EDs. We used the validated 4AT and care partner-completed AD8 screening tools, respectively, to exclude care partners of older adults with concern for delirium and include care partners of older adults with cognitive impairment. We conducted recorded, semi-structured interviews using a standardized guide, and two team members coded and analyzed all professional transcriptions to identify emerging themes and representative quotations.

**RESULTS:** Care partners’ mean age was 56.7 years, 80% were female, and 24% identified as African American. We identified four major barriers regarding ED discharge care transitions among care partners of PLWCI: 1) unique care considerations while in the ED setting impact the perceived success of the care transition, 2) poor communication and lack of care partner engagement was a commonplace during the ED discharge process, 3) care partners experienced challenges and additional responsibilities when aiding during acute illness and recovery phases, and 4) navigating the health care system after an ED encounter was perceived as difficult by care partners.

**DISCUSSION:** Our findings demonstrate critical barriers faced during ED discharge care transitions among care partners of PLWCI. Findings from this work may inform the development of novel care partner-reported outcome measures as well as ED discharge care transition interventions targeting care partners.

## 1. Introduction

Persons living with dementia (PLWD) and older adults with mild cognitive impairment (MCI) seek care in the emergency department (ED) at higher rates than their cognitively intact counterparts and account for nearly 2 million visits annually.^1-4^ Together, the majority of these persons living with cognitive impairment (PLWCI) are discharged from the ED,^3^ and their care partners subsequently provide a significant amount of hands-on care and navigation of health-related and social needs within this vulnerable time period. After an ED visit, PLWCI are at increased risk of adverse events, including functional decline, hospitalization, and mortality.^2,3,5-9^ The experiences of care partners have been more greatly described during the post-hospitalization care transition,^10-12^ yet little is known from the care partner’s perspective regarding the ED care transition.

With the ED visit often recognized as a sentinel event for older adults, efforts to improve care transitions for care partners of PLWCI have increased in recent years. ED discharge care transitions have been prioritized within the 2014 Geriatric ED Guidelines^13^ and the 2018 launch of the American College of Emergency Physicians geriatric ED accreditation process.^14^ Recent ED-centric initiatives, as part of the Geriatric Emergency care Applied Research 2.0 Network – Advancing Dementia Care,^15^ identified the need for the development of patient- and care partner-reported outcome measures that capture what matters most to stakeholders during ED care transitions.^6,16^

As a first step towards developing a novel care partner-reported outcome measure, we sought to assess the barriers experienced by care partners of PLWCI during ED discharge care transitions. A greater understanding of the ED discharge care transition, as experienced by care partners of PLWCI, will provide new insights into ways in which the ED discharge process can be improved. Additionally, this work will inform the subsequent development of tailored interventions or educational tools to support this population during ED discharge care transitions.

## 2. Methods

### 2.1 Study design

This study protocol was approved by the Yale University institutional review board. Methods and results are reported in accordance with the COnsolidated criteria for REporting Qualitative research (COREQ).^17^

### 2.2 Setting and participants

In this work, we chose to focus on care partners of PLWCI, as the unique experiences of those with intact cognition would likely identify separate barriers and themes during the ED discharge care transition. Described in greater detail below, PLWCI were identified through the electronic medical record as well as through in-ED cognitive testing to additionally capture those with MCI. Historically, the presence of dementia or MCI has been underdocumented in the electronic medical record and underrecognized in the ED.^18-20^ With evidence existing that these phenomena particularly impact racial and ethnic minorities, we performed cognitive testing of potential participants with validated tools to overcome limitations of prior research that potentially neglected to incorporate the perspectives of these groups despite higher rates of cognitive impairment.^21-23^

We recruited participants from four hospital EDs: a Level I trauma center/tertiary referral hospital, two academic community hospitals, and a freestanding ED within the same health system. Inclusion criteria for care partners were: 1) family member or friend of an ED patient aged 65 or older with impaired cognition, 2) anticipated discharge of the patient after the ED encounter, and 3) fluent in English or Spanish. To increase generalizability of our findings, patients could be community-dwelling or residing in any type of extended care facility at the time of ED visit. Exclusion criteria included: 1) intact cognition of the patient, 2) evidence of delirium, or 3) determination by the treating ED clinician that the candidate patient was inappropriate for the study due to critical illness, contact precautions, acute altered mental status, and intoxication among other reasons.

### 2.3 Data collection

I.U., a trained research assistant, approached and screened patients aged 65 years or older while in the ED, with purposive sampling by age, race, and chief complaint to capture a wide range of perspectives. I.U. assessed capacity and obtained informed consent (assent and proxy consent as indicated) in accordance with best research practices for the involved population and collected sociodemographic and contact information.^24,25^ Each care partner participant received a $25 gift card after study participation.

We performed cognitive assessments of all consenting older adults using the validated 4AT tool and the care partner-completed AD8 tool to serve as a proxy in identifying older adults with undiagnosed or undocumented cognitive impairment.^26-30^ The 4AT is scored from 0-12 and tests for cognitive impairment, including delirium. A score of ≥4 suggests possible delirium (prompting exclusion), a score of 1-3 suggests possible cognitive impairment, and a score of 0 suggests delirium or severe cognitive impairment is unlikely. Due to COVID-19 ED visitation policy restrictions, we then performed the care partner-completed AD8 by telephone with the family member or friend care partner contact person listed in the electronic medical record. Previously assessed in the ED,^26,27^ the AD8 is a screening tool more specifically focused on dementia with 8 questions and a score range of 0 to 8. Aligned with prior research,^28^ a score of ≥2 on the care partner-completed AD8 was noted to suggest cognitive impairment of the older adult patient and resulted in study inclusion.

K.H.B., a trained qualitative interviewer of care partners, contacted participants by telephone to complete one-on-one interviews within 3-7 days of the initial ED visit. This timeframe was chosen to mitigate recall bias while also allowing for a reasonable time period to elapse for the care partner to experience the ED transition and any associated barriers. We then developed a semi-structured interview guide based on the existing literature, our study team’s content expertise, and a conceptual model of ED care transitions among care partners of PLWCI. The conceptual model considered: 1) how upstream clinical and sociodemographic factors, as well as access to care considerations may lead a PLWCI to seek ED care, and 2) how ED care characteristics may impact subsequent discharge care transition outcomes of PLWCI as well as care partners. The semi-structured interview guide was then pilot tested and revised. All interviews were audio-recorded, professionally transcribed, and redacted. Field notes were taken immediately after each interview.

### 2.4 Data analysis

We used an iterative process of thematic analysis to synthesize the data, identify patterns, and develop themes across the interviews.^31^ Specifically, we used a combined deductive and inductive approach,^32^ where concepts in the standardized interview guide were used to structure the initial codebook and data from interviews were incorporated (Supplemental Text S1). The coding team was composed of C.J.G. and P.T.S., two emergency physicians with expertise in ED care transitions. C.J.G. has completed formal training in qualitative research. Initially, the two investigators independently coded five transcripts and met to review coding and refine the coding structure. Both researchers coded 100% of the data. Coders obtained consensus on major topics and subtopics from the remainder of the research team and applied the final coding structure to all transcripts. C.J.G. and P.T.S. iteratively reviewed coded data, compiled separate memos, and identified themes using NVivo software (version 12; QSR International, Victoria, Australia). Recruitment, interviewing, and coding occurred concurrently until data saturation was reached. Coders shared findings and obtained team consensus of representative quotes and contextualized findings.

## 3. Results

We approached 148 older adults in the ED between November 2021 and February 2022, and ultimately interviewed 25 care partners of PLWCI. On average, interviews lasted 29 minutes, with a range of 16 to 47 minutes. Care partner participants’ mean age was 56.7 years (standard deviation [SD] = 13.2 years), 80% were female, and 24% identified as African American. Participants frequently carried formal designations regarding the care of their family member or friend and reported a wide range of hours of caring per day. The patients cared for by the care partners resided primarily in the community (52%) and an almost even split was achieved among those with formal electronic medical record documentation of dementia and those with undocumented MCI. (Table 1)

**Table 1.** Sample characteristics (N = 25)

| **Variable** | **n (%)** |
| --- | --- |
| *Patient Characteristics* |  |
| Gender |  |
| Male | 7 (28) |
| Female | 18 (72) |
| Age, mean years (SD) | 83.2 (7.6) |
| Race |  |
| White | 19 (76) |
| African American | 6 (24) |
| Marital Status |  |
| Married | 9 (36) |
| Single | 7 (28) |
| Divorced | 3 (12) |
| Widowed | 6 (24) |
| Living Environment |  |
| Community | 13 (52) |
| Assisted Living Facility | 6 (24) |
| Skilled Nursing Facility | 6 (24) |
| ED Chief Complaint Category |  |
| Cardiopulmonary | 1 (4) |
| Musculoskeletal | 12 (48) |
| Gastrointestinal | 3 (12) |
| Neurologic | 4 (16) |
| Other | 5 (20) |
| Electronic medical record documentation of dementia | 12 (48) |
| *Care Partner Characteristics* |  |
| Designations (not mutually exclusive) |  |
| Legally Authorized Representative | 17 (68) |
| Surrogate Decision-Maker | 19 (76) |
| Healthcare Proxy | 19 (76) |
| Relation to the Patient |  |
| Grandchild | 1 (4) |
| Child | 16 (64) |
| Spouse | 4 (16) |
| Friend | 3 (12) |
| Sibling | 1 (4) |
| Hours Caring per Day |  |
| 1-4 | 10 |
| 5-8 | 1 |
| 8-12 | 0 |
| 13-23 | 3 |
| 24 | 8 |
| Not reported | 3 |
| Meets NASEM Caregiver definition | 25 (100) |
| Age, mean years (SD) | 56.7 (13.2) |
| Gender |  |
| Male | 5 (20) |
| Female | 20 (80) |
| Race |  |
| White | 19 (76) |
| African American | 6 (24) |
| Education |  |
| High School | 6 (24) |
| Some College | 3 (12) |
| Completed College | 6 (24) |
| Professional | 9 (36) |
| Not reported | 1 (4) |
| Care partner-completed AD8, mean (SD) | 6.7 (1.7) |

Four themes emerged: 1) unique care considerations while in the ED setting impact the perceived success of the care transition, 2) poor communication and lack of care partner engagement was a commonplace during the ED discharge process, 3) care partners experienced challenges and additional responsibilities when aiding during acute illness and recovery phases, and 4) navigating the health care system after an ED encounter was perceived as difficult by care partners. Themes, subthemes, and representative quotations of care partners are presented in Tables 2 and 3.

**Table 2.** Summary of Themes 1 & 2 and representative quotations

| Theme/Subtheme | Representative Quotation |
| --- | --- |
| **1. Unique care considerations while in the ED setting impact the perceived success of the care transition** |  |
| Change from familiar environment | I think patients with dementia that come into the emergency room require a whole different level of attention because their reactions to things, their confusion, their change in environment, all of that can have real impactful effects on them. (CP 186) |
| COVID-19 visitation policies | I know there's no visitors allowed, but when you have a patient in this condition [with dementia], it's not really a visitor. It's their caregiver. Recognizing that, yes, the nurses are doing that, and the doctors are doing their function, but no one knows the patient as well as the caregiver. I could tell that my mother was afraid. (CP 175) |
| Coordination of placement | Interviewer: What were some of your hopes for that emergency room visit? Interviewee: To help her find placement for any assisted living. We tried to deal with at-home nursing and stuff, and we weren't getting nursing fast enough for her. We were not getting the adequate help that she needed quick enough. We kept getting denied from the state and all this nonsense and all the runarounds. (CP 62) |
| Potential downplaying of cognitive impairment | ...her going in alone and people start talking to her like she's all there. She puts on a pretty good front until you talk to her long enough, you'll hear something that makes you go, "Wait a minute." I don't know what they're asking. I don't know what she's answering. (CP 185) |
| **2. Poor communication and lack of care partner engagement was a commonplace during the ED discharge process** |  |
| Competing priorities of emergency clinicians | We never saw the doctor [at discharge] because the trauma came in. The doctor went to the trauma, so when the nurse came over, the nurse says, "I'm so sorry, but she's kind of busy." The nurse went to speak with the doctor, and I guess the doctor must have said, "Yeah, print out the discharge instructions. The patient can go home." (CP 124) |
| Explanation of completed testing | I want somebody in layman terms to say, "We did this test. This was fine. We did this test. That concerned us." (CP 146) |
| Lengthy discharge paperwork | The pack of papers is getting thicker and thicker. Basically, all I need to know is that, did we change the medications? (CP 192) |
| Perceived premature decision for discharge | When it comes to Alzheimer's patients, they need to pay attention more because they do ask the memory questions and all that, but there's a lot more to it. I think they make too quick of a decision to release people and then there's more issues at home. (CP 62) |
| Communication with facility | The concept of discharging an 80-year-old dementia patient with discharge papers to go back to her rehab without anyone, any communication with anyone—who knows what happens? There's just no communication, and that feels like a tragedy waiting to happen. (CP 162) |
| Explicit instructions for next steps often lacking | For EDs, clarity about what they've done, what they've found. Any information they have about how she should proceed, like does she need bedrest for two days. Does she need a change in her medication? (CP 185) |
| Desire for a follow-up phone call | We never got a call back to say, "Hey, this is what the resolution was. We think it's X, Y, and Z, or we think it's nothing or whatever." I think that would be one of my serious considerations that maybe needs tightening up a little bit maybe, is some more communication with the family. (CP |
|  | 146) |

**Table 3.** Summary of Themes 3 & 4 and representative quotations

| Theme/Subtheme | Representative Quotation |
| --- | --- |
| **3. Care partners experienced challenges and additional responsibilities when aiding during acute illness and recovery phases** |  |
| Feelings of guilt | When I called them to let them know that she fell, broke her nose, and I'm blaming myself, and they're, "Stop. Stop. It's not your fault. You're doing the best job ever." They're my cheerleaders, my brothers. I just felt so awful because I didn't walk her to the car door. (CP 160) |
| Reliance on co-workers | I'm a teacher...I am going to miss a midterm tomorrow, but one of my colleagues is covering for me who teaches the same subject. (CP 178) |
| Feelings of obligation | [My mother, an additional caregiver] gets away at least a couple of hours but not as she should. It almost feels like if you're leaving your child home with someone while you run to the store and you feel that urgency to get home so you can take care of your child because you don't want to leave them too long...It's just that you just feel obligated to be there. (CP 82) |
| Making a best guess | That's part of the challenge too, is we don't know what she's experiencing because it's hard for her to verbalize it. (CP 146) |
| Enhancing in-home safety | Then I just went yesterday, myself and a carpenter friend of mine, we installed an aluminum railing system on the back there to help him so that it doesn't happen again. (CP 180) |
| Assistance from a broader social network | Then I thank God I have daughters that are willing to help because I still have to work to make ends meet. (CP 173) |
| Increased assistance with ADLs | I'm helping her more as far as total care. I'm assisting her more in washing her up and making sure she's dry and lotioning her body. Her walking is not the same as it was before so she's needing more assistance now.(CP 209) |
| **4. Navigating the health care system after an ED encounter was perceived as difficult by care partners** |  |
| Desire for community resource facilitation | For someone like myself to be able to have a resource or to be able to be directed to a resource where I can ask questions about the long-term care and/or how to get physical therapy. How do I get her some other assistance? Where can I just call and ask the question, is this normal, or is this really off the wall? That would be a great thing. (CP 175) |
| Primary care coordination | They always say, "Follow up with the primary care doctor and make an appointment with the primary care doctor." No one coordinates anything. She comes home, and it's a vicious circle. I'm out there flopping my arms in the wind. (CP 192) |
| Effort involved in completing follow-up appointments | Because she's immobile, we need to arrange for the van to take them out to appointments and such, so it's not as easy as call the doctor, make an appointment, and come tomorrow and check her out kind of thing. (CP 146) |
| Lack of health record interoperability | The problem with Dr. XXX is he's not a XXX [large integrated health care system] doc, so he doesn't get into the whole Epic thing...puts him behind the eight ball a little bit. (CP 146) |
| Degree of paperwork | They [Agency on Aging] have a boatload of paperwork that had to be filled out. I don't know what drives me more nuts, her anxiety or all this paperwork and red tape that I have to fill out. (CP 192) |
| Formal testing | I would love to be able to get her actually tested to see if she has Alzheimer's. I don't know where to look for that kind of stuff. (CP 147) |
| Respite care | Oh, yeah, I'm 24/7 [caregiver]. I really don't get any help. I did try and call |
|  | places to see if I can just get him for a couple of hours a week. I think it would be good for him and good for me, because he has a lot of anxiety. It's like having a five year old. They said that they don't have anything available. They had an extensive wait list. (CP 150) |
| Changing facility level of care | I have no confidence that she wasn't going to fall again, none, none at all. To me, it was—my next action was to find a 24-hour process to get her into and it was just a live-in place to get her out of there. I didn't think she would last very long there. (CP 162) |

### 3.1 Unique considerations in the ED setting

Care partners reported key aspects of acute care unique to the ED setting. Whether residing in the community or an extended care facility, care partners noted that PLWCI fare better in familiar environments and that the chaotic and potentially intense environment of the ED was particularly unsettling and distressing. Many care partners reported how this factor was amplified during the COVID-19 pandemic given limitations to visitor policies, exemplified by the following quotations:

> *I think that if you have patients like this [with dementia], certainly any change in the ordinary routine is very upsetting. The extra stimulation of being in one of the beds next to the nursing station out in a hallway, I think would be way too much for any of them to handle. (CP 175)*
>
> *It’s scary for that person. They want somebody there that they know. I just think that’s the most important thing that I would say any emergency hospital should allow that, even now with the COVID. (CP 179)*

Several care partners of those with MCI expressed desire for greater efforts regarding coordination of services, referrals, and placement from the ED. Care partners reported feeling like they were not getting adequate services (e.g. personal care or skilled care) in a timely fashion, and they believed the clinicians in the ED could be instrumental in helping them navigate their needs.

Care partners also reported that ED clinicians could seemingly be given a false sense of security by PLWCI, particularly if the patient intentionally downplayed existing cognitive impairment. Several care partners felt like this downplaying could hinder the initial stages of the cognitive evaluation process. One care partner noted:

> *They released my mom without talking to me at all. It’s because she told them that she was fine. That’s why I say I don’t think they asked her any questions to see if she had an issue with memory*…*I just really wish that she was tested for the confusion. My mother probably played it off so that they didn’t even think to do any specific tests. (CP 132)*

### 3.2 ED discharge process

Many care partners reported that the ED discharge process was substantially lacking in communication from staff and they did not feel they were adequately engaged during this key portion of the care transition. One care partner who was able to be present in the ED reported that the ED clinician had competing interests that prevented a more substantive one-on-one conversation at discharge, noting:

> *We never saw the doctor [at discharge] because the trauma came in. The doctor went to the trauma, so when the nurse came over, the nurse says, “I’m so sorry, but she’s kinda busy*.*” [The nurse] printed the instructions and we were able to go home. (CP 124)*

Collectively, care partners reported feeling there was a lack of explanation regarding management of the acute illness and anticipatory guidance. One care partner emphasized this sentiment by stating:

> *I think that there could have been some more instructions that said, “Okay, [your mother] is home. We found this. We think you should do X, Y, and Z*.*” Just maybe some more concise instructions that would help the family and the care partners and the aides so that we’re doing things based on recommendations of medical folks, not just what we’re watching on TV. (CP 146)*

ED encounters for PLWCI were also perceived to result in discharge deemed premature by several care partners. One care partner reported feeling be ‘kicked out’ by stating:

> *I would say it [ED visit] shouldn’t be like we’re kicking her out, and it’s your, now, responsibility, which is the impression that I got. Not just throwing her out on the curb. If you know that she has a problem with the bath—why are you sending her home until you understand how the bathroom and everything is related to her passing out? I shouldn’t be the detective to figure this one out. (CP 192)*

Care partners almost universally expressed desire for some type of clinician contact after an ED encounter, with one succinctly stating:

> *A call 24 hours later to find out how things are going would be really amazing. (CP 162)*

### 3.3 Care partner challenges and responsibilities

After an ED visit for a PLWCI, care partners experienced several challenges throughout the remainder of the acute illness and recovery phases. Prominently, care partners expressed the heightened need for close monitoring and follow-up after an ED visit and the difficulties met when balancing that with their professional responsibilities. One care partner stated:

> *Well, I work full time and I take care of their house. I live with him and I take care of him, so now I as a caregiver have a lot on me. First off, I’m working full time and 90 percent of the time I got my door closed and I’m talking to a doctor or somebody or trying to get some answers for my dad. As a caregiver, I need to know what to expect. It’s putting a lot of stress on me as to what do I have to do for him. (CP 201)*

Second, care partners often relied on a broad social network to assist in the post-ED care of PLWCI on very short notice. This included co-worker colleagues as well as grandchildren, with one care partner noting the impact the ED visit and care transition had on a young family member:

> *I have a 19-year-old daughter who goes to college. She was home for those three or four weeks between Christmas and New Year’s. I was crying. When she’s home, she helps me out a lot. (CP 192)*

Finally, care partners also highlighted significant ongoing responsibilities (e.g bill payment, house chores) related to the family member or friend that were exacerbated during the acute illness prompting ED visitation. These responsibilities were present even for care partners of a PLWCI residing in extended care facilities and were even more pronounced for care partners not living in the immediate vicinity of PLWCI.

### 3.4 Navigating the health care system

During the care transition period after an ED visit, care partners of PLWCI experienced difficulty in following up with primary care clinicians, obtaining desired information from community organizations regarding assistance, and changing the level of care at extended care facilities to increase monitoring during the recovery phase. One care partner who recognized a need for increased monitoring after the ED visit noted significant uncertainty, stating:

> *I find discharging to be a laughable process. It’s like, “All right. Well, you need 24-hour care*.*” Like, “Well, where does materialize from?” (CP 185)*

Care partners also expressed a lack of knowledge as to where to obtain formal neurocognitive testing, particularly for those with MCI. One care partner reported the impact that the recognized but undiagnosed status of their family member’s cognitive impairment had regarding facility placement decisions:

> *I need them to diagnose, prescribe, do what they need to do before any other stuff takes place because we’d like to keep her at home as long as possible. I’m not thinking institution, but I have to think of safety first because I can’t be there 24 hours a day. I still have to work. I don’t live there. I can’t say, “You can live with me*.*” (CP 173)*

Several care partners noted their continuous care for the patient needing emergency care, its impact on both care partner and the PLWCI, and their desire for information regarding options such as respite care. One care partner reported:

*I really would like to get him into some sort of daytime program…where it would be good for him and just where I know I can make an appointment and not have to worry about getting someone to check in on him. (CP 150)*

## 4. Discussion

This qualitative study revealed major barriers and care needs experienced by care partners of PLWCI navigating the ED discharge care transition. These barriers included unique care considerations in the ED setting that impact the perceived success of the care transition; poor communication and lack of care partner engagement during the ED discharge process; challenges experienced during the acute illness and recovery phases; and difficulty navigating the health care system after an ED encounter. These findings have implications for ED discharge discussions and for the development of ED care transition interventions targeting identified barriers among care partners of PLWCI.

Our work builds upon existing literature by identifying unique barriers experienced by care partners of PLWCI experiencing ED discharge care transitions. Prior qualitative research has assessed hospital staff training programs addressing PLWD,^33^ the role of the care partners in the ED itself,^34^ and cognitively intact older adults experiencing ED care transitions.^35^ Within the first theme presented, several care partners of PLWCI noted the chaotic and intense ED environment with amplification experienced with existing COVID-19 visitation policies. Despite the absence of evidence suggesting that visitation policies curbed COVID-19 transmission, 93% of hospitals in a nationally representative sample had a visitor restriction policy that extended to the ED, with only 39% of those hospitals having exceptions for persons with cognitive impairment.^36^ This work underscores the importance of such simple patient protections. As part of the second theme, care partners also noted the limitations of the ED discharge process, including a lack of explanation of testing, results, and next steps. In addressing ED discharge comprehension, available literature has largely excluded patients with cognitive impairment and/or their care partners,^37,38^ representing a potential area for future investigation to improve ED discharge care transitions for this population.

We additionally identified that care partners of PLWCI experienced several noteworthy challenges, largely centering on navigating the health care system during the ED discharge care transition. Our findings related to this critical barrier provide potential underlying etiologies as to why older adults with documented dementia experience greater 30-day ED revisit rates and other adverse outcomes compared to those without dementia.^2^ Identified within this work, care partners of PLWCI reported experiencing deficiencies in community resource access, primary care follow-up, respite care access, and completion of formal neurocognitive testing that may contribute to previously noted adverse outcomes. We therefore suggest that ED clinicians and investigators should consider these needs of care partners when discharging PLWCI. Spending extra time in the way of care coordination, case management, or referral facilitation in the ED could be instrumental in preventing care transition barriers commonly experienced by care partners.

Findings from this work will next be used to generate candidate items for inclusion within a novel care partner-outcome measure, specifically designed to incorporate care partner stakeholder priorities during ED care transitions. An ED-specific care partner-reported outcome measure has the potential to overcome limitations of currently available quality measures or tools due to their current absence of ED setting-specific considerations, inconsistent psychometric data, and deficiency-focused approaches to the care partner role.^39^ Furthermore, a newly-developed care partner-outcome measure will have the ability to quantitatively assess the ‘success’ of an ED discharge care transition from the perspective of a care partner, particularly having been developed based on their priorities and perspectives outlined in this work. Ultimately, prominent themes and subthemes identified within this work may also serve as a central component in developing interventions that improve care partner-reported outcome measures during ED discharge care transitions. To date, few ED-based interventions have been implemented with varying levels of success and have almost exclusively assessed healthcare utilization outcomes as opposed to care partner-reported outcome measures.^40-42^

### 4.1 Limitations

Our study findings should be considered in the context of several limitations. This research occurred in four EDs within one health system in New England, and findings may therefore not be generalizable to other regions. The study was conducted during the COVID-19 pandemic. It is possible that unique pandemic stressors and resource limitations led to suboptimal processes of care than would have occurred otherwise. To enhance generalizability, we purposively enrolled participants to ensure a diverse sample by age, race, and chief complaint category. However, we were unable to enroll any participants that identified Spanish as their preferred language despite intentional efforts to translate study consent forms and materials and the presence of bilingual and bicultural study team members. We recognize that inclusion of a greater proportion of non-English-speaking participants may reveal additional barriers and themes surrounding ED care transitions.

## 5. Conclusion

Our findings demonstrate critical barriers faced during ED discharge care transitions among care partners of PLWCI. Findings from this work may inform the development of novel care partner-reported outcome measures as well as ED discharge care transition interventions targeting care partners.

## Data Availability

All data produced in the present study are available upon reasonable request to the authors.

## Acknowledgments & Disclosure of Funding

Dr. Gettel is a Pepper Scholar with support from the Claude D. Pepper Older Americans Independence Center at Yale School of Medicine (P30AG021342), the National Institute on Aging (NIA) of the National Institutes of Health (R03AG073988), the Alzheimer’s Association (ARCOM-22-878456), and the NIA Imbedded Pragmatic Alzheimer’s and AD-Related Dementias Clinical Trials Collaboratory (NIA IMPACT Collaboratory; U54AG063546). Dr. Venkatesh is supported by the American Board of Emergency Medicine National Academy of Medicine Anniversary fellowship and previously by the Yale Center for Clinical Investigation (KL2TR000140) from the National Center for Advancing Translational Science. Dr. Cohen is supported by the NIA (K76AG059987). Dr. Monin is supported by the NIA (U54AG063546, R01AG058565). Dr. Feder received funding from the National Heart Lung and Blood Institute (K12HL138037) of the NIH and the Yale Center for Implementation Science. Dr. Hwang is supported by the NIA (R33AG058926, R61AG069822), by the John A Hartford Foundation, and the West Health Institute. The funders had no role in the design and conduct of the study; collection, management, analysis, and interpretation of the data; and preparation or approval of the manuscript.

## References

1. Centers for Disease Control and Prevention. National Hospital Ambulatory Medical Care Survey: 2019 Emergency Department Summary Tables. Accessed May 9, 2022. https://www.cdc.gov/nchs/data/nhamcs/web_tables/2019-nhamcs-ed-web-tables-508.pdf.

2. Kent T, Lesser A, Israni J, Hwang U, Carpenter C, Ko KJ. 30-day emergency department revisit rates among older adults with documented dementia. J Am Geriatr Soc. 2019;67(11):2254–2259.

3. LaMantia MA, Stump TE,Messina FC, Miller DK, Callahan CM. Emergency department use among older adults with dementia. Alzheimer Dis Assoc Disord. 2016;30(1):35–40.

4. Hunt LJ, Coombs LA, Stephens CE. Emergency department use by community-dwelling individuals with dementia in the United States: an integrative review. J Gerontol Nurs. 2018;44(3):23–30.

5. Lucke JA, de Gelder J, Heringhaus C, et al. Impaired cognition is associated with adverse outcomes in older patients in the Emergency Department; the Acutely Presenting Older Patients (APOP) study. Age Ageing. 2018;47(5):679–684.

6. Gettel CJ, Falvey JR, Gifford A, et al. Emergency department care transitions for patients with cognitive impairment: a scoping review. J Am Med Dir Assoc. 2022;S1525-8610(22)00154-2.

7. Feng Z, Coots LA, Kaganova Y, Wiener JM. Hospital and ED use among Medicare beneficiaries with dementia varies by setting and proximity to death. Health Aff (Millwood). 2014;33(4):683–690.

8. Stephens CE, Newcomer R, Blegen M, Miller B, Harrington C. The effects of cognitive impairment on nursing home residents’ emergency department visits and hospitalizations. Alzheimers Dement. 2014;10(6):835–843.

9. de Gelder J, Lucke JA, de Groot B, et al. Predictors and outcomes of revisits in older adults discharged from the emergency department. J Am Geriatr Soc. 2018;66(4):735–741.

10. Griffiths S, Stephen G, Kiran T, Okrainec K. “She knows best”: a qualitative study of patient and caregiver views on the role of the primary care physician follow-up post-hospital discharge in individuals admitted with chronic obstructive pulmonary disease or congestive heart failure. BMC Fam Pract. 2021;22(1):176.

11. Hahn-Goldberg S, Jeffs L, Troup A, Kubba R, Okrainec K. “We are doing it together”; the integral role of caregivers in a patients’ transition home from the medicine unit. PLoS One. 2018;13(5):e0197831.

12. Pritchard E, Cussen A, Delafosse V, Swift M, Jolliffe L, Yeates H. Interventions supporting caregiver readiness when caring for patients with dementia following discharge home: a mixed-methods systematic review. Australas J Ageing. 2020;39(3):e239–e250.

13. American College of Emergency Physicians, American Geriatrics Society, Emergency Nurses Association, Society for Academic Emergency Medicine, Geriatric Emergency Department Guidelines Task Force. Geriatric emergency department guidelines. Ann Emerg Med. 2014;63(5):e7–25.

14. Geriatric Emergency Department Accreditation Program. American College of Emergency Physicians. Accessed May 1, 2022. https://www.acep.org/geda/.

15. Hwang U, Carpenter C, Dresden S, et al. The Geriatric Emergency care Applied Research network approach: a protocol to advance stakeholder consensus and research priorities in geriatrics and dementia care in the emergency department. BMJ Open. 2022;12(4):e060974.

16. Gettel CJ, Voils CI, Bristol AA, et al. Care transitions and social needs: a Geriatric Emergency care Applied Research (GEAR) Network scoping review and consensus statement. Acad Emerg Med. 2021;28(12):1430–1439.

17. Tong A, Sainsbury P, Craig J. Consolidated criteria for reporting qualitative research (COREQ): a 32-item checklist for interviews and focus groups. Int J Qual Health Care. 2007;19:349–57.

18. Gilmore-Bykovskyi AL, Jin Y, Gleason C, et al. Recruitment and retention of underrepresented populations in Alzheimer’s disease research: a systematic review. Alzheimers Dement (N Y). 2019;5:751–770.

19. Quiñones AR, Mitchell SL, Jackson JD, et al. Achieving health equity in embedded pragmatic clinical trials for people living with dementia and their family caregivers. J Am Geriatr Soc. 2020;68 Suppl 2(Suppl 2):S8–S13.

20. Gleason CE, Norton D, Zuelsdorff M, et al. Association between enrollment factors and incident cognitive impairment in Blacks and Whites: data from the Alzheimer’s Disease Center. Alzheimers Dement. 2019;15(12):1533–1545.

21. Mayeda ER, Glymour MM, Quesenberry CP, Whitmer RA. Inequalities in dementia incidence between six racial and ethnic groups over 14□years. Alzheimers Dement. 2016;12(3):216–224.

22. Cooper C, Tandy AR, Balamurali TBS, Livingston G. A systematic review and meta-analysis of ethnic differences in use of dementia treatment, care, and research. Am J Geriatr Psychiatry. 2010;18(3):193–203.

23. Gilmore-Bykovskyi A, Johnson R, Walljasper L, Block L, Werner N. Underreporting of gender and race/ethnicity differences in NIH-funded dementia caregiver support interventions. Am J Alzheimers Dis Other Demen. 2018;33(3):145–152.

24. Prusaczyk B, Cherney SM, Carpenter CR, DuBois JM. Informed consent to research with cognitively impaired adults: transdisciplinary challenges and opportunities. Clin Gerontol. 2017;40(1):63–73.

25. Fields LM, Calvert JD. Informed consent procedures with cognitively impaired patients: a review of ethics and best practices. Psychiatry Clin Neurosci. 2015;69(8):462–471.

26. Carpenter CR, DesPain B, Keeling TN, Shah M, Rothenberger M. The six-item screener and AD8 for the detection of cognitive impairment in geriatric emergency department patients. Ann Emerg Med. 2011;57(6):653–661.

27. Carpenter CR, Bassett ER, Fischer GM, Shirshekan J, Galvin JE, Morris JC. Four sensitive screening tools to detect cognitive dysfunction in geriatric emergency department patients: Brief Alzheimer’s Screen, Short Blessed Test, Ottawa 3DY, and the caregiver-completed AD8. Acad Emerg Med. 2011;18(4):374–384.

28. Galvin JE, Roe CM, Coats MA, Morris JC. Patient’s rating of cognitive ability: using the AD8, a brief informant interview, as a self-rating tool to detect dementia. Arch Neurol. 2007;64(5):725–730.

29. O’Sullivan D, Brady N, Manning E, et al. Validation of the 6-item Cognitive Impairment Test and the 4AT test for combined delirium and dementia screening in older emergency department attendees. Age Ageing. 2018;47(1):61–68.

30. Tieges Z, MacLullich AJM, Anand A, et al. Diagnostic accuracy of the 4AT for delirium detection in older adults: systematic review and meta-analysis. Age Ageing. 2021;50(3):733–743.

31. Kiger ME, Varpio L. Thematic analysis of qualitative data: AMEE guide no. 131. Med Teach. 2020;42(8):846–854.

32. Fereday J, Muir-Cochrane E. Demonstrating rigor using thematic analysis: a hybrid approach of inductive and deductive coding and theme development. Int J Qual Methods. 2006;5(1):80–92.

33. Schneider J, Miller J, Teschauer W, Kruse A, Teichmann B. Evaluation and effectiveness of a two-day dementia training program for hospital staff working in an emergency department. J Alzheimers Dis. 2021;84(4):1634–1644.

34. Fry M, Elliott R, Murphy S, Curtis K. The role and contribution of family carers accompanying community-living older people with cognitive impairment to the emergency department: an interview study. J Clin Nurs. 2022;31(7-8):975–984.

35. Gettel CJ, Hayes K, Shield RR, Guthrie KM, Goldberg EM. Care transition decisions after a fall-related emergency department visit: a qualitative study of patients’ and caregivers’ experiences. Acad Emerg Med. 2020;27(9):876–886.

36. Lo AX, Wedel LK, Liu SW, et al. COVID-19 hospital and emergency department visitor policies in the United States: impact on persons with cognitive or physical impairment or receiving end-of-life care. J Am Coll Emerg Physicians Open. 2022;3(1):e12622.

37. Sheikh H, Brezar A, Dzwonek A, Yau L, Calder LA. Patient understanding of discharge instructions in the emergency department: do different patients need different approaches? Int J Emerg Med. 2018;11(1):5.

38. McCarthy DM, Buckley BA, Engel KG, Forth VE, Adams JG, Cameron KA. Understanding patient-provider conversations: what are we talking about? Acad Emerg Med. 2013;20(5):441–448.

39. Hanson LC, Bennett AV, Jonsson M, et al. Selecting outcomes to ensure pragmatic trials are relevant to people living with dementia. J Am Geriatr Soc. 2020;68(Suppl 2):S55–S61.

40. Shah MN, Jacobsohn GC, McJones C, et al. Care transitions intervention reduces ED revisits in cognitively impaired patients. Alzheimers Dement (N Y). 2022;8(1):e12261.

41. Gettel CJ, Goldberg EM. Fall prevention intervention in the emergency department for older adults. J Aging Sci. 2020;8(1):222.

42. Shaw FE, Bond J, Richardson DA, et al. Multifactorial intervention after a fall in older people with cognitive impairment and dementia presenting to the accident and emergency department: randomised controlled trial. BMJ. 2003;326(7380):73.

