## Supplemental Text S1 for "Emergency department care transition barriers: a qualitative study of care partners of older adults with cognitive impairment"

**Supplemental Text S1. Semi-structured patient interview guide**

We are developing a caregiver-reported outcome measure for caregivers of older adults with cognitive impairment experiencing ED discharge care transitions. We recognize that your family member or loved one may or may not have a diagnosis of dementia, but memory problems were noted based on some of the questions from the ED the other day. We would like to get your perspective about their care and present condition and the role that you play. We also want to understand what you as the family member (or caregiver) experience after your loved one left the ED, allowing clinicians to better provide you care and address expectations while you and your loved one are in the ED. This interview will take approximately 30-45 minutes, depending on how much time you have to share your perspectives. What you say will never be identified with you specifically, and there are no right or wrong answers. We will share a summary of the study results with you. With your permission, I will be audio recording the conversation. Do you have any questions or concerns before we get started?

1. Icebreaker: How are you related to the older adult enrolled in the ED? How long have you known them? Have you noticed memory problems with them? If so, what specifically have you noticed? How long have you been involved with their medical or daily care – if at all? Tell me about your loved one’s visit to the ED. What brought them there? As a family member or caregiver, what were your hopes for the visit? Were you in touch with a primary care provider prior to seeking care in the ED?
2. Discharge Process: What happened when your loved one was discharged from the ED? Can you tell me about the process? Were you contacted? What were you told was causing your loved one’s symptoms? What instructions were you given? Were you provided information on when to seek care again if needed?
3. Care Transition: What happened in the days after your loved one returned home after the ED visit? Can you walk we through those days? Did you talk with/see their primary care provider? Did any questions come up during that time period, and if so, how did you get them addressed? Did you have family to help you during this time? What challenges did you face during this time? When thinking about caring for your loved one, what do you think would help you most?
4. Caregiver: Describe how you obtain fulfillment in your role as family member or caregiver. Does anything specific bring you joy or happiness in taking care of them or helping them? Have you ever found that caring for your loved one has been stressful? Has your quality of life changed through your loved one’s illness? Do you feel like your physical or mental health has suffered as a result of your loved one’s illness and the requirements in caring for them just after the ED visit and over the longer term? Do you feel that because of the time you spend with your relative that you don’t have enough time for yourself? What coping and adaptive strategies have you used? Have you faced out-of-pocket healthcare expenditures? Have you had to miss work or been less productive as a result of your loved one’s illness? Have you had to make changes to childcare? Do you have anyone else in the family or as part of a network of people that helps you with responsibilities in caring for your loved one? Is there anything else you would like to share about your role as a caregiver as it relates to after-ED care?
5. Expectations: Is there anything that you expected to happen in the ED or days after that did not? Why do you think that might be?
